# A Balanced Bagging Classifier machine learning model-based web application to predict risk of febrile neutropenia in cancer patients

**DOI:** 10.1101/2024.08.06.24311562

**Authors:** HŞ Bozcuk, M. Yıldız

## Abstract

**Background:** Although several models exist to predict risk of febrile neutropenia in cancer patients, there is still need to more accurately quantify this risk to minimize morbidity of and mortality from this treatment toxicity.

**Material and methods:** From previous reports of our group, un updated predictive model had emerged. We refined our algorithm even further by using Balanced Bagging Classifier (BBC) machine learning in the previous model derivation cohort, discarding all the missing data. Moreover, we made a web application to make it accessible for experimental clinical use.

**Results:** We used clinical data from 3439 cycles of chemotherapy obtained from the periods of 2010-2011 and 2015-2019, with 133 episodes of febrile neutropenia observed (after 4% of chemotherapy cycles). BBC resulted in a more efficient model as reflected by an area under curve (AUC) of 0.97, accuracy of 0.95, sensitivity of 0.93, and specificity of 0.95. Permutation importance analysis revealed previous febrile neutropenia, cancer type and receipt of previous radiotherapy as the most important features for the BBC model. The web app that integrates the BBC model with a user-friendly user interface has been found to be clinically useful.

**Conclusions:** Using machine learning with our previous data, we are now able to predict the risk of febrile neutropenia more effectively after chemotherapy in cancer patients. The resultant web application is functional and makes use of the developed machine learning model to predict febrile neutropenia.

## INTRODUCTION

Febrile neutropenia (FN) remains a critical concern in oncology, characterized by a fever and a significant reduction in neutrophil count, typically induced by chemotherapy. It poses a life-threatening risk to patients, requiring immediate medical intervention (1). The accurate prediction of FN risk is crucial for optimizing prophylactic measures and patient management strategies. Traditional predictive models have leveraged clinical factors such as age, type of cancer, chemotherapy regimen, and baseline blood counts to estimate FN risk (2,3). In 2 previous publications from our group, we were able to delineate the predictors of febrile neutropenia in cancer (4,5). However, our models and also models from other groups lacked precision at times, mostly with low sensitivity potentially leading to mostly under-, or sometimes, overprediction of the risk of febrile neutropenia.

Machine learning (ML) approaches have increasingly been applied in healthcare to enhance predictive accuracy (6). Techniques such as random forests, support vector machines, and neural networks have demonstrated success in various medical predictions (6-8). A significant challenge in developing ML models for FN prediction lies in the imbalanced nature of clinical datasets, where the occurrence of FN is relatively rare compared to non-FN events. For example, the proportion of cases within the whole cohort was 1.5% and 4% in our previous model derivation cohorts (4,5). This imbalance can lead to models biased towards predicting the majority class, thereby underperforming in identifying FN cases.

The Balanced Bagging Classifier (BBC) is an ensemble learning technique specifically designed to address the issue of class imbalance (9). By resampling the training data, BBC aims to balance the class distribution, thereby improving the model’s sensitivity and specificity for the minority class. This study builds on our previous work to refine an existing predictive model for FN by incorporating BBC, which has been shown to enhance the model’s ability to accurately predict FN risk.

In addition to developing a robust predictive model, we have aimed to test the feasibility of a web application to facilitate its use in clinical settings. This application potentially provides healthcare providers with an accessible tool to estimate FN risk, aiding in the decision-making process for patient care.

## MATERIALS AND METHODS

### General

Data had already been collected and the details of the cohorts had been published (4,5). The combined derivation cohort from these publications were carefully examined, and all chemotherapy cycles with at least one feature missing were eliminated. So, the resultant remaining dataset included all cycles and cases from the combined derivation datasets without any missing data from the periods of 2010-2011 and 2015-2019. The resultant dataset included demographic information, clinical features, and treatment details. Of these cycles, 133 resulted in FN, representing approximately 4% of the total cycles. Key variables included patient age, cancer type, chemotherapy regimen, prior FN episodes, previous radiotherapy, and other relevant clinical, treatment and precycle laboratory factors. The dataset was then split into training (75%) and test (25%) sets. The training set was used for model development, while the test set served to evaluate the model’s performance.

### Model Development

The BBC was employed to address the class imbalance problem. BBC is an ensemble method that constructs multiple base classifiers trained on balanced subsets of the data (10,11). This method reduces the bias towards the majority class, improving the model’s ability to predict FN. Refer to Figure 1 for a demonstration of the working principles of BBC.

**Figure 1.**
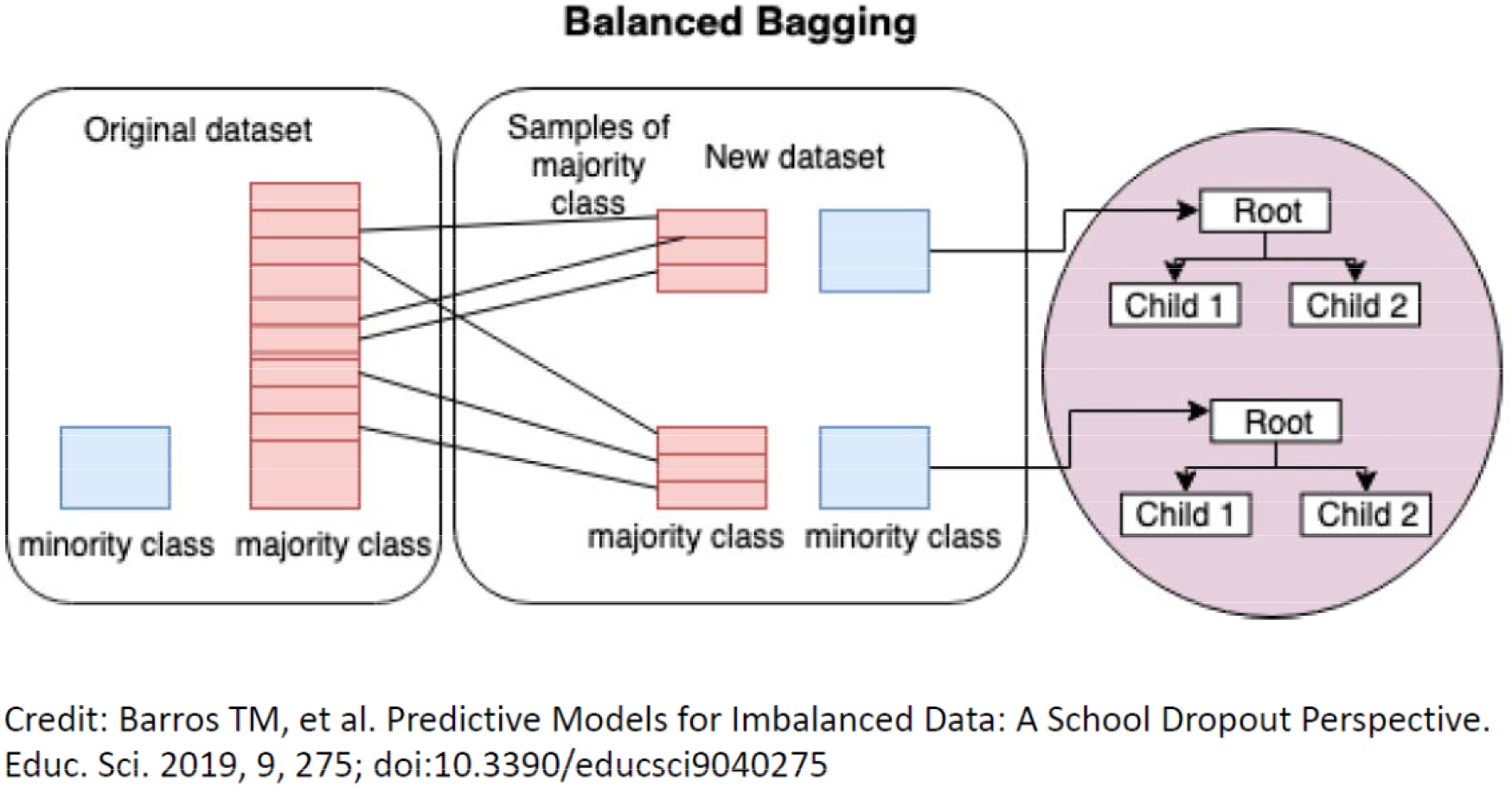
Balanced Bagging Classifier (BBC)

Then, in order to identify the most influential features in the model, we used permutation importance analysis (12). The permutation importance algorithm involves computing importance i_j_ for feature f_j_, computing the score s_kj_ for each feature j and for each repetition k, as follows:

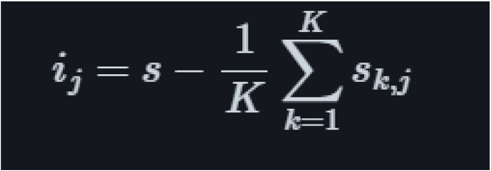

The model’s parameters were optimized using cross-validation, which helps prevent overfitting and ensures the model generalizes well to unseen data. We evaluated the model’s performance using metrics such as the area under the receiver operating characteristic curve (AUC), accuracy, sensitivity, and specificity. The AUC provides a measure of the model’s discriminative power, while accuracy, sensitivity, and specificity offer insights into its overall performance and its ability to correctly identify FN and non-FN cases.

As a part of the analysis, we searched optimal value from a grid. To achieve this, we found the probabilities for the class label first, then looked for the optimal threshold to map the probabilities to its proper class label. The probability of prediction can be obtained from a classifier by using predict_proba() method from the Sklearn Python library (13). Using the optimal threshold, then we calculated the confusion matrix, and the accuracy, sensitivity and specificity figures from the sklearn. metrics module.

### Web Application Development

The resultant model file was saved by the help of Pickle Python library, and the model file and the accompanying files were loaded to a GitHub repository, and then was deployed using the Streamlit Python library (14,15). The web application enabled the entry of model predictors before a cycle of chemotherapy and then calculation of the risk of febrile neutropenia after a cycle of chemotherapy.

## RESULTS

### General

A total of 3439 chemotherapy cycles were examined. The most common diagnosis was breast cancer (38%), and 4% of the cycles ended with febrile neutropenia. Median cycle number on current protocol was 3. See Table 1 for the characteristics of chemotherapy cycles and related features.

**Table 1.** Chemotherapy cycles and related features.

| Features | n | % | Median | Mean | Standard deviation |
| --- | --- | --- | --- | --- | --- |
| Chemotherapy cycles | 3439 | 100 |  |  |  |
| *General* |  |  |  |  |  |
| Cancer type |  |  |  |  |  |
| Breast cancer | 1315 | 38 |  |  |  |
| Lung cancer | 890 | 26 |  |  |  |
| Colorectal cancer | 1159 | 34 |  |  |  |
| Other cancers | 75 | 2 |  |  |  |
| Stage |  |  |  |  |  |
| 1 to 3 | 1927 |  |  |  |  |
| 4 | 1512 |  |  |  |  |
| Involved systems number* |  |  | 0 | 0.8 | 1.1 |
| Gender |  |  |  |  |  |
| Female | 1811 | 53 |  |  |  |
| Male | 1628 | 47 |  |  |  |
| Age |  |  | 55 | 55.1 | 11.4 |
| ECOG performance status |  |  | 1 | 0.7 | 0.7 |
| Coronary disease |  |  |  |  |  |
| No | 3133 | 91 |  |  |  |
| Yes | 306 | 9 |  |  |  |
| Chronic obstructive Lung disease (COLD) |  |  |  |  |  |
| No | 3252 | 95 |  |  |  |
| Yes | 187 | 5 |  |  |  |
| Radiotherapy |  |  |  |  |  |
| Did not receive before | 3372 | 98 |  |  |  |
| Received before | 67 | 2 |  |  |  |
| Previous chemotherapy |  |  |  |  |  |
| No | 1922 | 56 |  |  |  |
| Yes | 1517 | 44 |  |  |  |
| *Treatment* |  |  |  |  |  |
| Treatment as an inpatient |  |  |  |  |  |
| No | 3240 | 94 |  |  |  |
| Yes | 199 | 6 |  |  |  |
| CSF usage |  |  |  |  |  |
| No | 2543 | 74 |  |  |  |
| Yes | 896 | 26 |  |  |  |
| Chemotherapy dose reduction |  |  |  |  |  |
| No | 3117 | 91 |  |  |  |
| Yes | 322 | 9 |  |  |  |
| Previous febrile neutropenia |  |  |  |  |  |
| No | 3211 | 93 |  |  |  |
| Yes | 228 | 7 |  |  |  |
| Febrile neutropenia after chemotherapy |  |  |  |  |  |
| No | 3306 | 96 |  |  |  |
| Yes | 133 | 4 |  |  |  |
| Drug number** |  |  |  |  |  |
| 1 | 336 | 10 |  |  |  |
| 2 | 1644 | 48 |  |  |  |
| 3 | 1076 | 31 |  |  |  |
| 4 | 382 | 11 |  |  |  |
| 5 | 1 | 0 |  |  |  |
| Regimen risk*** |  |  |  |  |  |
| 1 | 938 | 27 |  |  |  |
| 2 | 2157 | 63 |  |  |  |
| 3 | 344 | 10 |  |  |  |
| Cycle no on current protocol |  |  | 3 | 2.8 | 1.4 |
| *Laboratory* |  |  |  |  |  |
| LDH (IU/ml) |  |  | 343 | 370 | 235 |
| ALT (IU/ml) |  |  | 18 | 22 | 20 |
| Creatinine (mg/dl) |  |  | 0.71 | 0.76 | 0.22 |
| Lymphocyte count (x1000/mm3) |  |  | 1.7 | 1.9 | 1.3 |
| Albumin (mg/dl) |  |  | 4.2 | 4.2 | 0.4 |

\*, Number of metastatic organ systems, an example: if there are 8 lung and 3 liver metastases, and no other metastases, number of metastatic organ systems is 2 (lung and liver), \*\*, total number of drugs in the treatment protocol including chemotherapy, biological and targeted agents, \*\*\*; Baseline febrile neutropenia risk classification for the current chemotherapy protocol according to NCCN

### BBC model

The BBC model achieved an AUC of 0.97, indicating the discriminative ability. The model’s accuracy was 0.95, with a sensitivity of 0.93 and a specificity of 0.95. These results were related to the model’s effectiveness in terms of correctly identifying both FN and non-FN cases, to judge its potential utility in clinical practice. Refer to Figure 2 for the AUC plot.

**Figure 2.**
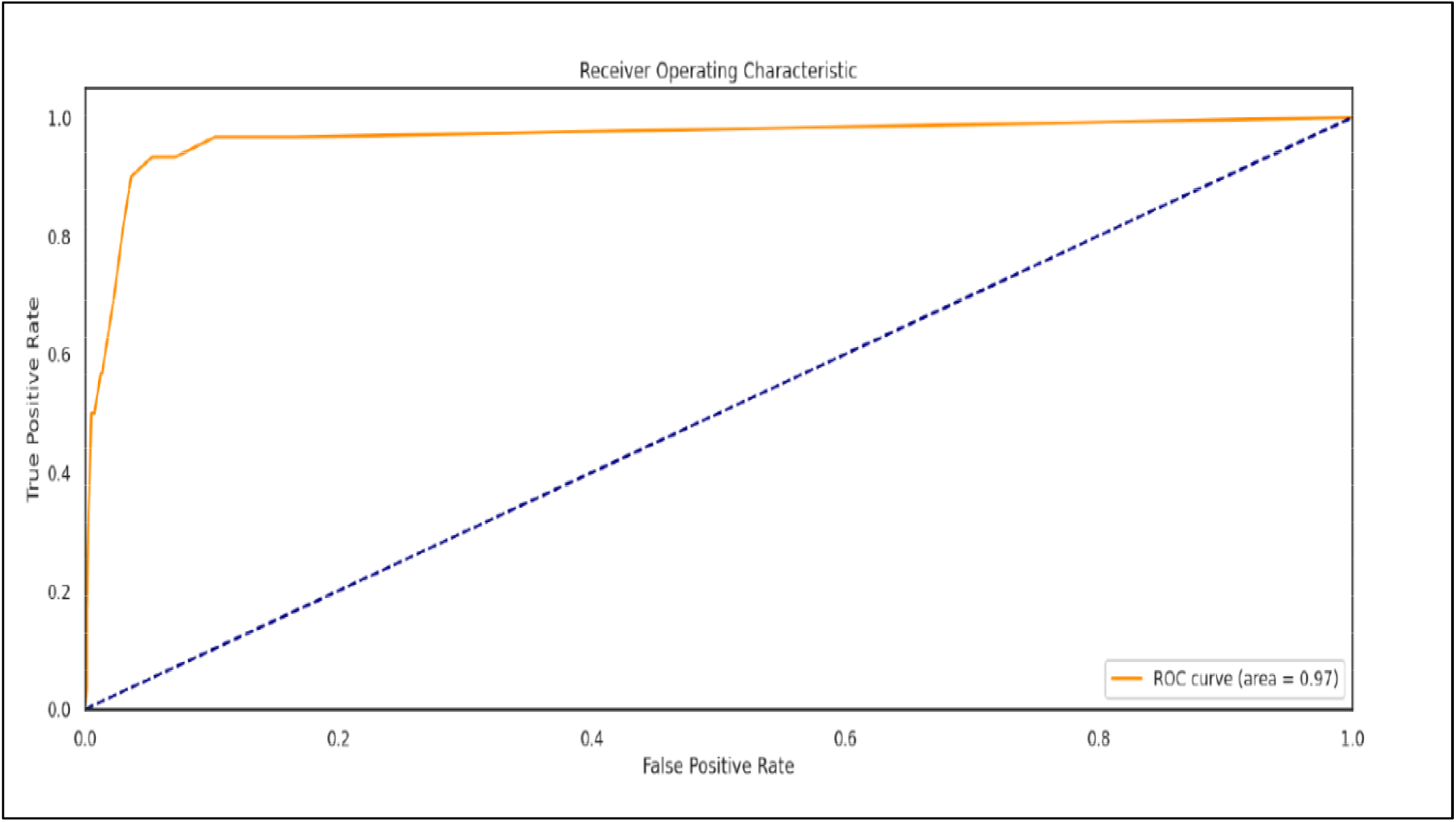
The AUC plot

Permutation importance analysis further highlighted the significance of specific clinical factors. Refer to Figure 3 for the results of the permutation importance analysis. Notably, patients with a history of FN were at a higher risk of experiencing FN during subsequent chemotherapy cycles. Cancer type also played a crucial role, with certain cancers associated with a higher risk of FN. Previous radiotherapy was another significant factor, that correlated with the risk of febrile neutropenia after chemotherapy.

**Figure 3.**
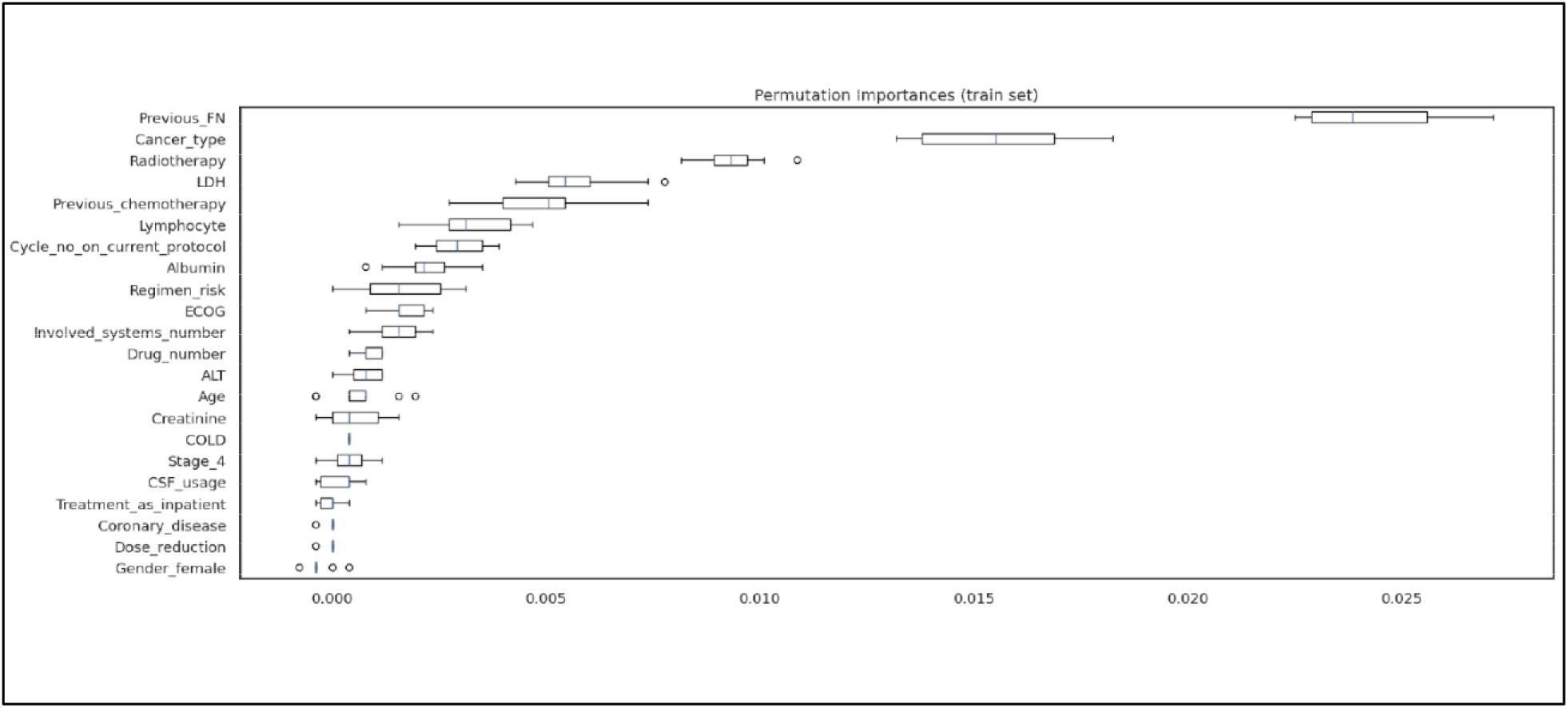
Permutation importance analysis

Three most influential factors from the BBC model can be viewed in Figure 4. These features are displayed in the plot with reference to the magnitude of their importance scores. Figure 4a focuses on the effect of previous febrile neutropenia, 4b on cancer type and 4c on radiotherapy history, in relation to febrile neutropenia risk.

**Figure 4.**
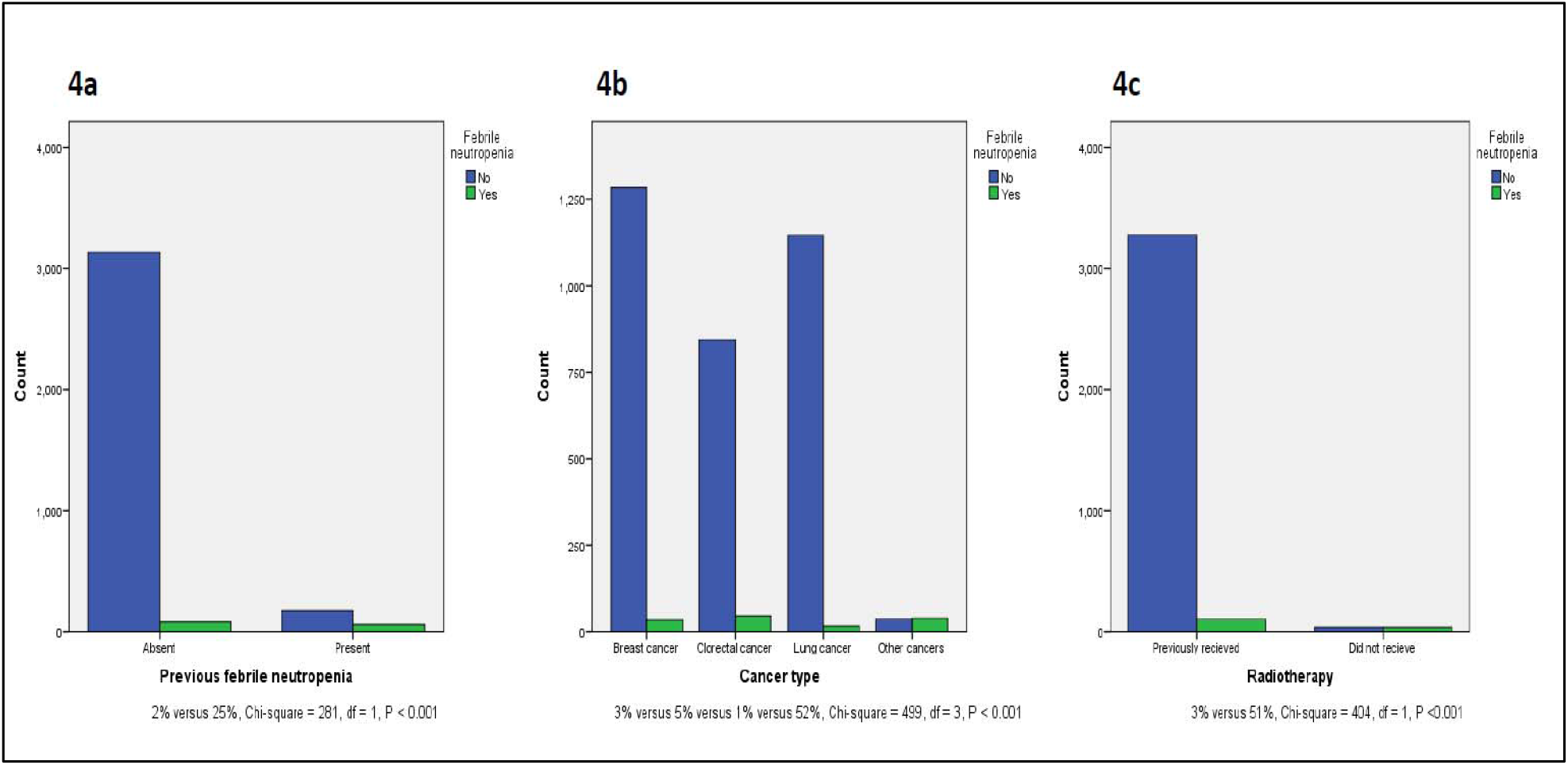
Most influential factors on the risk of febrile neutropenia

### Web application

The web application developed as part of this study integrates the BBC model and provides a user-friendly interface for clinical use. It allows healthcare providers to input patient data and receive real-time predictions of FN risk after a specific cycle of chemotherapy. The application was evaluated for usability and found to be a practical tool in clinical settings. Web application user interfaces can be viewed in Figure 5. Figure 5a reflects the data entry part, and Figure 5b details the febrile neutropenia risk output part.

**Figure 5.**
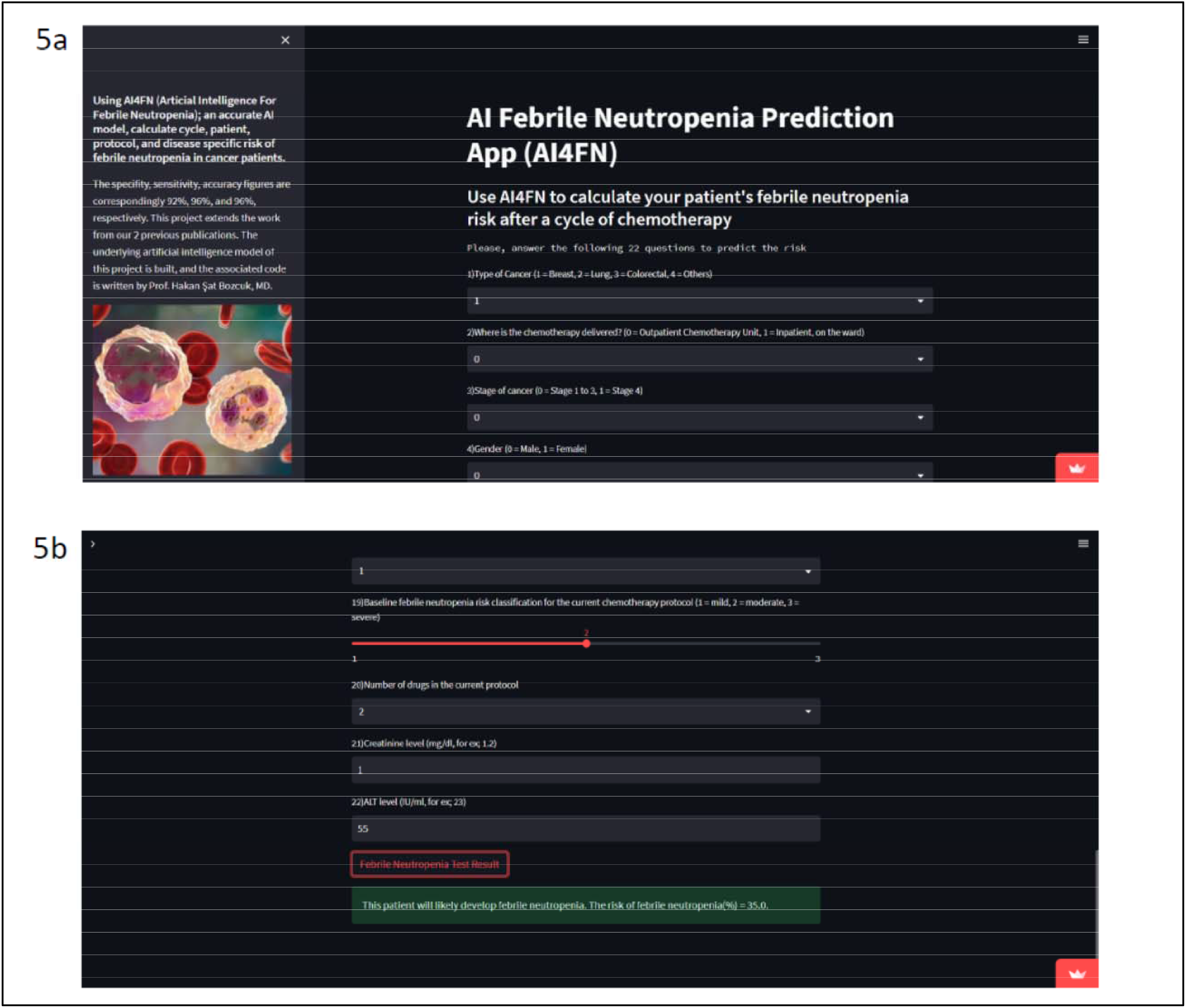
Web application user interface

## CONCLUSION

The implementation of the Balanced Bagging Classifier (BBC) in our study has yielded an accurate predictive model for febrile neutropenia (FN) in cancer patients undergoing chemotherapy. The model’s notable performance metrics, including an AUC of 0.97, accuracy of 0.95, sensitivity of 0.93, and specificity of 0.95, highlight its potential utility in clinical practice. These findings are consistent with existing literature that emphasizes the capability of ensemble learning methods in enhancing predictive accuracy, especially in the context of imbalanced clinical datasets. BBC, as an ensemble learning method, involves creating multiple versions of a predictor and using these to get an aggregated prediction. Specifically, BBC aims to address class imbalance by training each base classifier on balanced subsets of the data, which can improve the model’s sensitivity to the minority class. This method is particularly useful when the dataset contains a significantly lower number of instances of one class compared to another, as it helps prevent the model from becoming biased toward the majority class (16,17).

The high sensitivity and specificity observed in our model suggest that it is well-calibrated to distinguish between FN and non-FN cases. This accuracy is crucial in a clinical setting, where the consequences of both false positives and false negatives can be significant. For instance, a false positive could lead to unnecessary prophylactic interventions, which can be costly and potentially harmful due to side effects like myalgia, while a false negative could result in inadequate preparation for an FN episode, putting the patient at serious risk and may lead to complications (18,19).

The permutation importance analysis provided valuable insights into the most significant predictors of FN risk, with previous FN episodes, cancer type, and prior radiotherapy identified as the most influential factors. The strong association of previous FN episodes with subsequent risk is well-documented, suggesting that a history of FN is a critical marker for assessing patient vulnerability. Similarly, the type of cancer plays a pivotal role, with non-small cell lung cancer and small cell lung cancer, for example, being associated with higher FN risk due to the intensive nature of their treatment regimens. Prior radiotherapy’s contribution to FN risk may be linked to its cumulative effect on bone marrow suppression, which can be exacerbated by concurrent chemotherapy (20-23).

Our study also demonstrated the practical application of the BBC model through the development of a web application, which provides a user-friendly interface for healthcare providers. This tool allows for real-time FN risk assessment, facilitating timely and informed clinical decisions. The importance of integrating predictive models into clinical workflows has been highlighted in recent studies, which emphasize the need for accessible and interpretable tools to support medical professionals (24). The usability testing of our application suggests that it could be effectively implemented in routine clinical practice, potentially improving patient outcomes by enabling early intervention and tailored prophylaxis.

The success of our model aligns with the broader trend in healthcare towards the adoption of machine learning and artificial intelligence (AI) technologies (25). The ability of these technologies to process vast amounts of data and identify complex patterns offers unprecedented opportunities for personalized medicine. Recent advances in AI have shown promise in various domains, from diagnostic imaging to treatment planning, underscoring the transformative potential of these technologies (26). However, our study is not without limitations. The exclusion of missing data, while necessary to maintain model integrity, may introduce bias and limit the generalizability of our findings. Missing data is a common issue in clinical datasets, and strategies such as imputation or the use of advanced models that can handle missingness should be considered in future research (27). Additionally, while our model demonstrated high accuracy on the test set, its real-world applicability needs to be validated through prospective clinical trials. Such validation is crucial to ensure that the model’s performance translates effectively into practical clinical settings.

The promising results of this study suggest several avenues for future research. One potential direction is the incorporation of additional clinical and molecular markers, which could further enhance the model’s predictive power. For instance, genetic polymorphisms related to drug metabolism or immune response could provide additional layers of information, helping to identify patients at the highest risk of FN (28). Another area of interest is the exploration of model interpretability. As machine learning models become increasingly complex, ensuring that their predictions are interpretable and transparent is essential for clinical acceptance and trust.

In conclusion, our study demonstrates the efficacy of the Balanced Bagging Classifier in predicting febrile neutropenia risk in cancer patients undergoing chemotherapy. The model’s integration into a user-friendly web application represents a significant step towards practical clinical implementation, offering a valuable tool for febrile neutropenia risk assessment and management. From a broader point of view, as the field of AI in healthcare continues to evolve, it is imperative to explore and address the challenges and opportunities that these technologies present, including our model, as we highlighted in this study.

## Data Availability

All data produced in the present study are available upon reasonable request to the authors

## REFERENCES

1- Boccia R, Glaspy J, Crawford J, Aapro M. Chemotherapy-Induced Neutropenia and Febrile Neutropenia in the US: A Beast of Burden That Needs to Be Tamed? Oncologist, 2022;27(8):625–636. doi: 10.1093/oncolo/oyac074.

2- Lyman GH, Lyman CH, Agboola O. Risk Models for Predicting Chemotherapy-Induced Neutropenia. Oncologist, 2005;10(6):427–437. 10.1634/theoncologist.10-6-427.

3- Ba Y, Shi Y, Jiang W, et al. Current management of chemotherapy-induced neutropenia in adults: key points and new challenges. Cancer Biol Med, 2020. doi: 10.20892/j.issn.2095-3941.2020.0069

4- Bozcuk H, Yildiz M, Artaç M, et al. A prospectively validated nomogram for predicting the risk of chemotherapy-induced febrile neutropenia: a multicenter study. Support Care Cancer, 2015; 23:1759– 1767. DOI 10.1007/s00520-014-2531-6.

5- Bozcuk H, Coşkun HŞ, İlhan Y, et al. Prospective external validation of an updated algorithm to quantify risk of febrile neutropenia in cancer patients after a cycle of chemotherapy. Supportive Care in Cancer, 2022; 30:2621–2629. 10.1007/s00520-021-06681-0.

6- An Q, Rahman S, Zhou J, Kang JJ. A comprehensive review on Machine Learning in healthcare industry: classification, restrictions, opportunities and challenges. Sensors (Basel), 2023; 23(9): 4178. doi: 10.3390/s23094178.

7- Islam MM, Haque MR, Iqbal H, et al. Breast cancer prediction: a comparative study using Machine Learning techniques. SN Computer Science, 2020; 1:290. 10.1007/s42979-020-00305-w.

8- Liu S, Du H, Feng M. Robust Predictive Models in Clinical Data—Random Forest and Support Vector Machines. In “Leveraging Data Science for Global Health”, 2020; 219–228.

9- Mashette N. Balanced Bagging Classifier (Bagging for Imbalanced Classification), 2023. Medium. At https://medium.com/@nageshmashette32/balanced-bagging-classifier-bagging-for-imbalanced-classification-dfba66c44c14. Accessed at 4.8.2024.

10- Bagging Classifier. Imbalanced Learn. At https://imbalanced-learn.org/stable/ensemble.html. Accessed at 4.8.2024.

11- BaggingClassifier. Scikit Learn. At https://scikit-learn.org/stable/modules/generated/sklearn.ensemble.BaggingClassifier.html. Accessed at 4.8.2024.

12- Permutation Importance. Learn Tutorial. At https://www.kaggle.com/code/dansbecker/permutation-importance. Accessed at 4.8.2024.

13- scikit-learn 1.5.1. At https://pypi.org/project/scikit-learn/. Accessed at 4.8.2024.

14- Ellis M. How to Use Python Pickle [+Examples]. At https://blog.hubspot.com/website/python-pickle. Accessed at 4.8.2024.

15- Streamlit. At https://github.com/streamlit/streamlit. Accessed at 4.8.2024.

16- Brownlee J. Bagging and Random Forest for Imbalanced Classification. Machine Learning Mastery. At https://machinelearningmastery.com/bagging-and-random-forest-for-imbalanced-classification/. Accessed at 4.8.2024.

17- Yevonnael A. Imbalanced Classification & Balanced Classifier. Kaggle. At https://www.kaggle.com/code/yevonnaelandrew/imbalanced-classification-balanced-classifier. Accessed at 4.8.2024.

18- Lapidari P, Vaz-Luiz I, Meglio AD. Side effects of using granulocyte-colony stimulating factors as prophylaxis of febrile neutropenia in cancer patients: A systematic review. Crit Rev Oncol Hematol, 2021; 157:103193. doi: 10.1016/j.critrevonc.2020.103193.

19- Punnapuzha S, Edemobi PK, Elmoheen A. Febrile Neutropenia. StatPearls. At https://www.ncbi.nlm.nih.gov/books/NBK541102/. Accessed at 4.8.2024.

20- Zatarah R, Faqeer N, Quraan T, et al. Validation of the FENCE risk groups for prediction of Febrile Neutropenia with first-cycle chemotherapy. JNCI Cancer Spectrum (2022) 6(3): pkac038. 10.1093/jncics/pkac038.

21- Flanigan JA, Yasuda M, Chen CC, Li EC. ChemotherapylZJinduced febrile neutropenia (FN): healthcare resource utilization (HCRU) and costs in commercially insured patients in the US. Supportive Care in Cancer, 2024;32:373. 10.1007/s00520-024-08492-5.

22- Lyman GH, Abella E, Pettengell R. Risk factors for febrile neutropenia among patients with cancer receiving chemotherapy: A systematic review. Critical Reviews in Oncology/Hematology, 2014; 90(3): 190–199.

23- Crespo A, Forbes L, Vu K, et al. Prevention and Outpatient Management of Febrile Neutropenia in Adult Cancer Patients. Clinical Practice Guideline. Ontario Health, 2021. At https://www.cancercareontario.ca/en/file/63491/download?token=F7OoHvFg. Accessed at 4.8.2024.

24- Rahmani AM, Yousefpoor E, Yousefpoor MS, et al. Machine Learning (ML) in Medicine: Review, Applications, and Challenges. Mathematics, 2021; 9(22): 2970. 10.3390/math9222970.

25- May M. Eight ways machine learning is assisting medicine. Nature Medicine, 2021; 27: 2–3.

26- Shehab M, Abualigah L, Shambour Q, et al. Machine learning in medical applications: A review of state-of-the-art methods. Computers in Biology and Medicine, 2022; 145: 105458. 10.1016/j.compbiomed.2022.105458.

27- Buuren SV. Flexible Imputation of Missing Data, Second Edition, 2018. Chapman and Hall/CRC. 10.1201/9780429492259.

28- Faraji A, Manshadi HRD, Mobaraki M, Zare M, Houshmand M. Association of ABCB1 and SLC22A16 Gene Polymorphisms with Incidence of Doxorubicin-Induced Febrile Neutropenia: A Survey of Iranian Breast Cancer Patients. PLoS One, 2016;11(12): e0168519. doi: 10.1371/journal.pone.0168519. eCollection 2016.

